# Personalized Risk-Prediction Tool for Deceased Donor Kidney Offers: Stakeholder Perspectives from a Qualitative Study

**DOI:** 10.64898/2026.03.02.26347468

**Authors:** Kelly Chong, Igor Litvinovich, Christos Argyropoulos, Rachel Taylor, Yiliang Zhu

**Affiliations:** Department of Internal Medicine, University of New Mexico Health Sciences Center; University of New Mexico, Southwest Center for Advancing Clinical and Translational Innovation, Albuquerque, NM, United States

**Keywords:** deceased donor kidney, decision support tool, kidney transplant risk, qualitative research, shared decision making

## Abstract

**Background and Hypothesis:** Kidney transplant (KT) candidates in the United States face prolonged wait times, while deceased donor kidney (DDK) discard rates continue to rise. Existing tools such as the Kidney Donor Profile Index (KDPI) provide largely population-level risk estimates and have limited utility for individualized decision making at the time of an offer. We sought stakeholder input on the content, design, and implementation of a prototype web-based Kidney Risk Calculator intended to support patient-centered decision making during DDK offer evaluation. We hypothesized that patients and providers would view risk and benefit estimates of KT outcomes, individualized to a donor–recipient pair, as a useful aid for kidney-offer decisions.

**Methods:** We conducted a qualitative study using focus groups and individual interviews with transplant stakeholders at a single transplant center. Participants included transplant candidates, a patient advocate, transplant coordinators, and transplant physicians. Semi-structured sessions included a live demonstration of the prototype app, after which participants provided feedback regarding usability, interpretability, contextual information needs, perceived clinical utility, and anticipated barriers and facilitators. Sessions were recorded, transcribed, and analyzed using inductive reflexive thematic analysis.

**Results:** Stakeholders viewed individualized outcome projections as a helpful adjunct to clinical judgment, particularly for higher-risk offers. Key design priorities included: (1) educational content on hepatitis C virus, Public Health Service risk criteria, calculated panel reactive antibody (cPRA), and dialysis-versus-transplant trade-offs; (2) plain-language narratives, simple visuals, minimal use of acronyms, U.S. customary units, and stepwise user input; and (3) alignment with time-sensitive, phone-based donor-offer workflows and variable levels of digital access.

**Conclusions:** Stakeholders highlighted the importance of combining individualized outcome predictions with accessible risk communication and integration into existing clinical workflows. These findings can inform the design, implementation, and evaluation of decision-support tools for kidney-offer discussions.

**Key learning points:** *What was known:* Kidney transplant risk tools, such as KDPI, aim to inform KT stakeholders of the quality of a DDK and associated average risks and benefits. However, little is known about stakeholder perspectives on tool features, usability, communication of individualized risk, or integration into clinical workflows.

*This study adds:* Perspectives of transplant candidates, coordinators, and clinicians regarding individualized donor–recipient risk prediction, their acceptance of the tool as a useful adjunct to clinical judgment, and their emphasis of clear communication, educational context, usability, and alignment with time-sensitive kidney-offer workflows.

*Potential impact:* These findings can inform the design, refinement, and deployment of kidney-offer decision-support tools by addressing implementation barriers related to communication, confidentiality, workflow constraints, and health and digital literacy.

## INTRODUCTION

Kidney transplantation (KT) is the preferred treatment for end-stage kidney disease (ESKD), offering superior survival, quality of life, and cost-effectiveness compared with dialysis.^1–3^ However, the supply of donor kidneys continues to fall short of demand in the U.S., and transplant programs in the U.S. increasingly consider deceased donor kidneys (DDK) from older donors or donors with comorbidities, often referred to as “marginal” or higher-risk organs.^4,5^ Nevertheless, DDKs that were procured for transplant but were discarded have increased from approximately 20% prior to the COVID-19 pandemic to nearly 29.3% in 2024,^6,7^ underscoring the need for effective risk communication to address uncertainties and concerns in kidney-offer acceptance, particularly for higher-risk donors.

Several factors contribute to DDK discards. Providers often face uncertainties about how donor characteristics interact with individual patient profiles; acceptance thresholds vary across providers and centers; and few tools exist to support efficient, personalized decision making when a DDK offer is received.^4^ Standard approaches such as the Kidney Donor Profile Index (KDPI) provide population-level risk assessments from a donor quality perspective, but have notable limitations for clinical decision making.^8^ KDPI is donor-focused, does not generate individualized survival estimates for a specific donor–recipient pair, and offers only modest discrimination for graft survival.^9^ Consequently, its utility for shared decision making is limited, especially when accepting a high-risk donor may benefit candidates who have had prolonged dialysis or limited future opportunities for transplantation.

To address these limitations, we developed a prototype web-based Kidney Risk Calculator app, a decision aid designed for use during kidney-offer evaluation.^9,10^ It integrates paired donor and recipient characteristics to generate personalized predictions of post-KT outcomes as well as projected outcomes associated with declining the offer and returning to the waitlist. In the proposed clinical workflow, donor and recipient characteristics are entered into the app at the time of organ offer, and the personalized outcome estimates are used in discussions between providers and candidates during offer evaluation.

Because transplant decision making is complex and time-sensitive, quantitative performance metrics alone cannot fully capture the factors that influence how decision-support tools are used in practice. Although decision-support tools for DDK transplantation and patient-centered approaches to their development have been described,^11,12^ important gaps remain in understanding how transplant stakeholders interpret individualized risk information, which features they find most useful, and how such tools can be implemented in clinical practice.

Guided by International Patient Decision Aid Standards (IPDAS) and user-centered design principles,^13^ we conducted a qualitative study involving transplant candidates, transplant coordinators, and transplant clinicians. Our objective was to elicit stakeholder perspectives on the app’s content, features, functionality, and anticipated barriers and facilitators to adoption. These insights are intended to guide future refinement of the prototype and support integration of similar tools into routine transplant care.

## MATERIALS AND METHODS

### Study design and setting

This study was approved by the University of New Mexico Human Research Protection Office (HRPO 23-071). We conducted focus groups and individual interviews at the University of New Mexico Hospital (UNMH) Kidney Transplant Center. This early-phase formative evaluation was conducted to obtain stakeholder feedback regarding the prototype Kidney Risk Calculator app.

### User-centered design framework

The Kidney Risk Calculator app was developed using an iterative, user-centered design approach. Early engagement of end users was prioritized to enhance relevance, clarity, and usability. Figure 1 depicts the design process, including stakeholder engagement, feedback analysis, and refinement.

**Figure 1.**
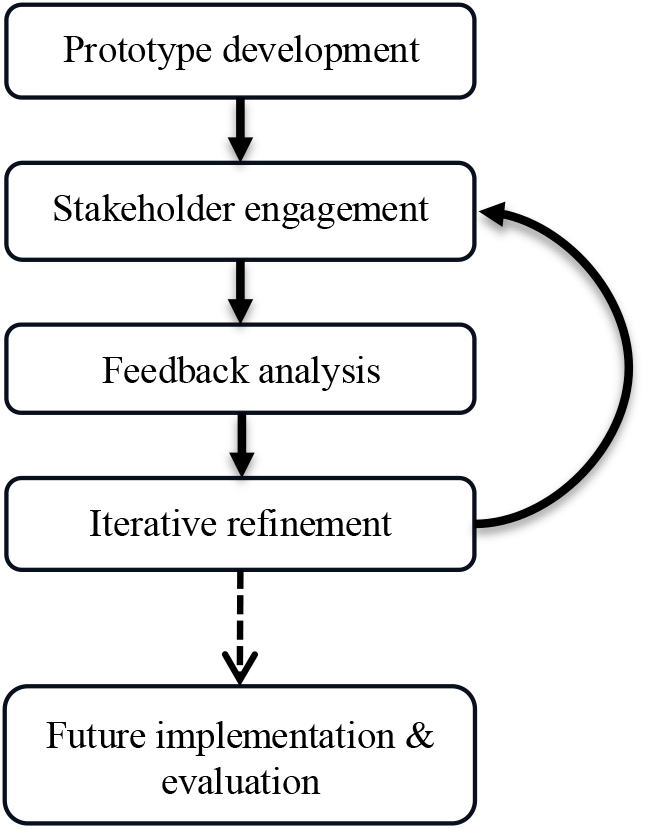
User-centered design process for the Kidney Risk Calculator app. Prototype development was followed by stakeholder engagement, feedback analysis, and iterative refinement.

### Participants and recruitment

We recruited three stakeholder groups from the UNMH Kidney Transplant Center: transplant providers, transplant coordinators, and transplant candidates, including one patient advocate. Because of scheduling constraints and limited provider availability, all five invited transplant providers completed individual interviews. Of eight coordinators invited, three participated in a focus group. Transplant candidates on the UNMH kidney transplant waitlist were first stratified by sex, race/ethnicity, and dialysis status and then randomly selected. Of 30 candidates invited by email, five participated in a focus group.

### Data collection

Between December 2022 and May 2023, we conducted focus groups and interviews via secure, password-protected Zoom videoconferencing sessions (Zoom Video Communications, Inc., San Jose, CA, USA). Each session began with a live demonstration of the prototype Kidney Risk Calculator app^10^ (Figures 2a & 2b) and lasted approximately 45 to 60 minutes.

**Figure 2a.**
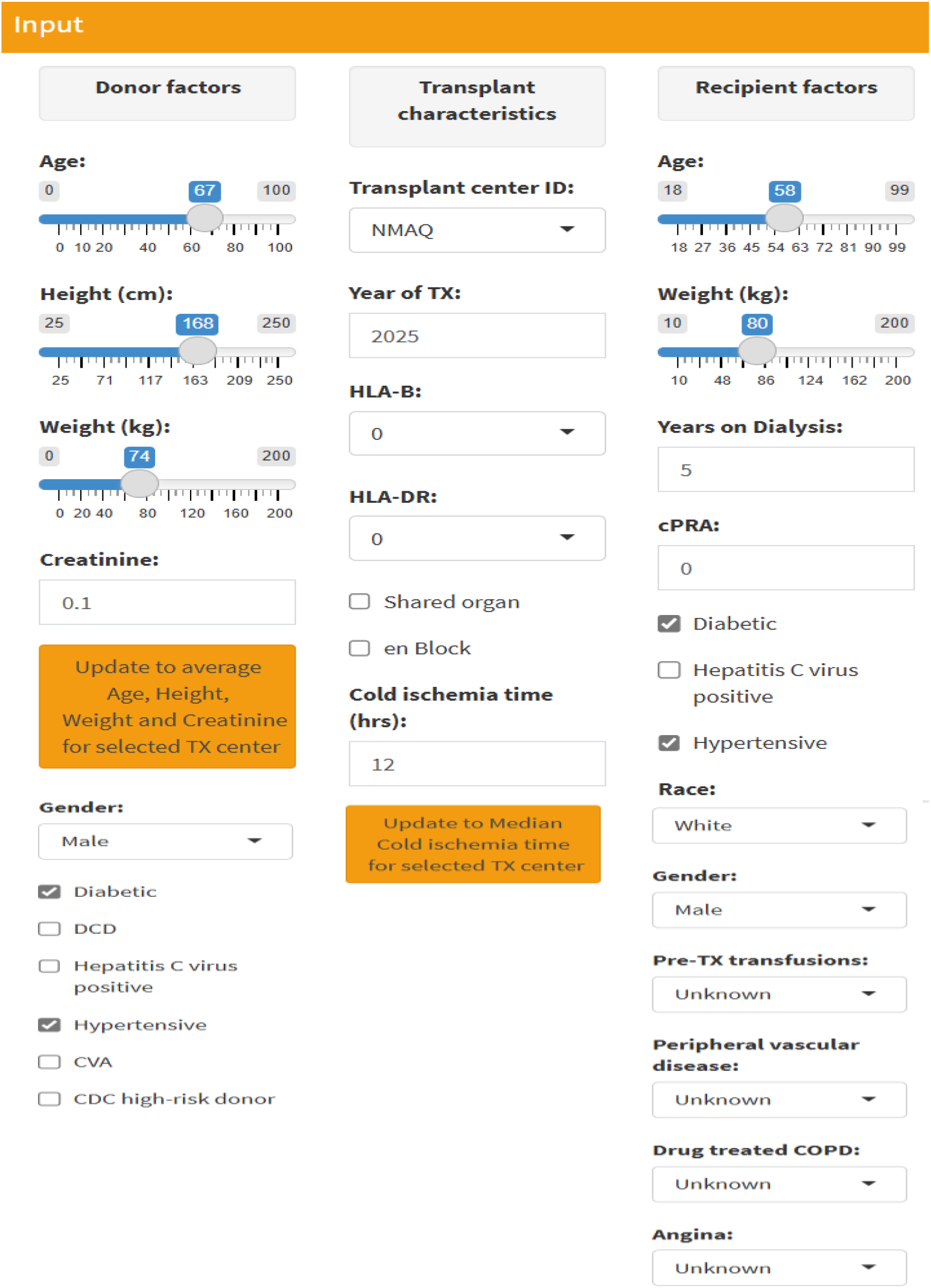
Kidney Risk Calculator Interface Input Layout. User input screen for entering donor, recipient, and transplant characteristics into the prototype Kidney Risk Calculator app.

**Figure 2b.**
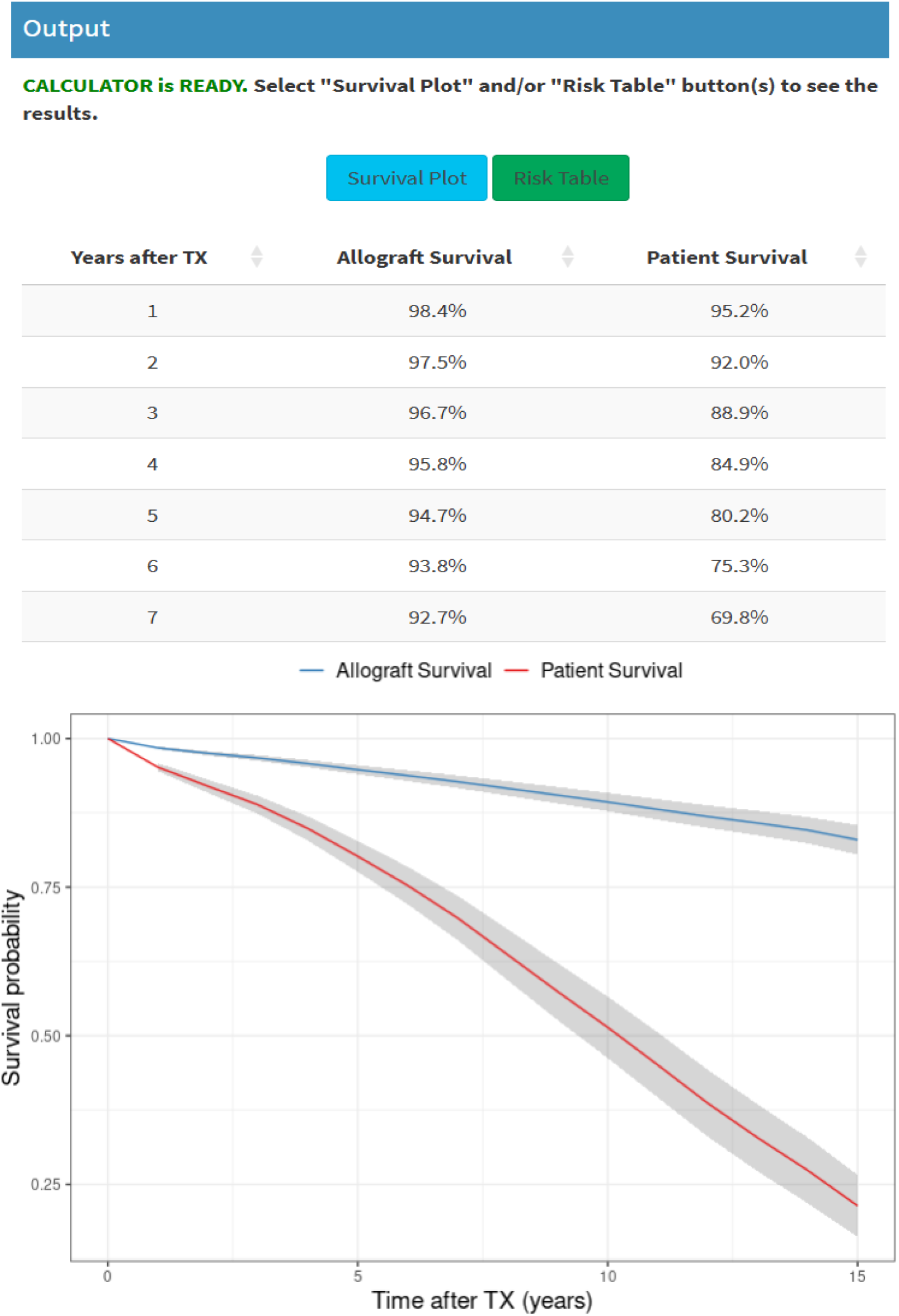
Kidney Risk Calculator Interface Output Layout. Output screen displaying projected post-transplant outcomes and projected outcomes associated with declining the offer and remaining on dialysis.

Participants provided feedback immediately following the demonstration. Using a semi-structured guide, we explored stakeholder perceptions of (1) app content (clarity, missing information, and relevance); (2) format and features (visual display, navigation, and usability); (3) functionality (perceived utility and workflow fit); and (4) anticipated barriers and facilitators. All sessions were recorded, professionally transcribed, and de-identified for analysis. The interview guide is provided in Supplementary Table S1.

### Data analysis

We conducted an exploratory, formative qualitative evaluation using inductive reflexive thematic analysis.^14,15^ Consistent with early-stage, user-centered design, we did not seek thematic saturation.^16^ Two qualitative research analysts independently reviewed transcripts, maintained reflexive memos, documented interpretive decisions, met for peer debriefing, and jointly developed the codebook. The primary analyst coded all transcripts in NVivo (QSR International, 2020),^17^ and the secondary analyst double-coded a subset to enrich interpretation. Inter-rater reliability was not calculated, consistent with reflexive thematic analysis.^14^ We conducted matrix queries to examine the distribution of coded references across stakeholder groups and thematic domains. A matrix summary table and descriptive heatmaps (Supplementary Table S2 and Supplementary Figures S1 and S2) are provided to illustrate cross-group representation and support quotation selection. The heatmap was used descriptively to illustrate the distribution of coded content across stakeholder groups and not to infer the relative importance of themes.

## RESULTS

### Participant demographics

A total of 13 stakeholders participated: 4 transplant candidates, 1 patient advocate, 3 transplant coordinators, and 5 transplant providers (3 nephrologists and 2 advanced practice providers). Among the five patient-stakeholder participants (four transplant candidates and one patient advocate), two were female and three were male; reported race/ethnicity included Black, Native American, and White. All coordinators were female, and providers included three females and two males. Race and ethnicity were not collected for coordinators or providers. Participant characteristics are summarized in Table 1.

**Table 1.** Participant Characteristics.

| Characteristic | Category | Patient Stakeholders<br>(n=5) | Coordinators<br>(n=3) | Providers<br>(n=5) |
| --- | --- | --- | --- | --- |
| Sex | Female | 2 | 3 | 3 |
|  | Male | 3 | 0 | 2 |
| Race/Ethnicity | Black | 1 |  |  |
|  | Native American | 1 |  |  |
|  | White | 2 |  |  |
|  | Unknown | 1 |  |  |

Race/ethnicity was collected for patient-stakeholder participants only; coordinators and providers participated in the study in their professional roles.

### Thematic findings

Analysis yielded inductively developed codes organized into three domains: content, format and features, and functionality. Some themes recurred across domains (e.g., education regarding hepatitis C virus [HCV] and workflow fit). Below, we present analytic summaries and illustrative quotations to reflect convergence and divergence across stakeholder groups.

#### 1) Content: Education on HCV, cPRA, and survival trade-offs; avoiding information overload; adding missing context

Patient stakeholders emphasized information about HCV, cPRA, and transplant outcomes as central to decision making, while cautioning against information overload. Several participants expressed a desire for concise educational content that could be revisited after clinic visits and presented in modular, plain-language sections.

> “…it would be helpful to have additional information about Hep-C risks and benefits using the app so patients can review the information beyond medical appointment conversations.”

Willingness to accept DDK varied; some patients prioritized access and timing of DDK availability over granular donor characteristics. When asked whether the app’s output would be sufficient to decide on donor acceptance within an hour, one participant stated, “I’ll take whatever they give me,” echoed by another’s “Heck yeah,” indicating that for some patients, donor availability outweighed specific aspects of kidney quality. Another participant similarly expressed willingness to accept “any kidney,” while remaining “super skeptical” about HCV.

Providers viewed the app’s incorporation of recipient factors as addressing a gap in KDPI-only assessments. One provider noted:

> “…this is new and different… we have the probability considering both the donor and the recipient, which we currently lack because we go by KDPIs and donors.”

Stakeholders suggested additional variables and contextual details to better align predicted outcomes with clinical practice and patient concerns, including coronary artery disease (rather than angina), diabetes management (A1c), dialysis adherence, smoking or substance use, kidney size, recent hospitalizations, cold ischemic time defaults (the preset cold ischemia time assumptions used when actual values are unavailable), and use of the last (rather than average) donor creatinine. Some providers questioned whether donor HCV status should meaningfully contribute to risk calculations in the direct-acting antiviral era:

> “…Hep C should probably not be included in the calculation… those kidneys actually do really well.”

Coordinators requested clearer education to contextualize “higher-risk donors,” specifically Public Health Service (PHS) risk criteria, to reduce refusals driven by misconceptions. They described patients often conflating “high-risk” with infection transmission despite negative testing, and recommended embedding explanations of how such designations are assigned:

> “…they automatically associate it with, ‘Oh, I’m going to get HIV.’”

Participants also identified content gaps not yet modeled, including COVID-positive donors. As one coordinator asked:

> “…how much would it change if we accepted a COVID-positive donor for this patient?
>
> … I don’t know how that would affect probabilities … because that’s not something we’re doing.”

They additionally emphasized the salience of time on dialysis. One patient noted that the app did not capture personal decline while waiting:

> “I’m running out of time… I’m going downhill pretty quick.”

#### 2) Format and features: Plain-language narratives, simple visuals, stepwise inputs, and U.S. customary units

Patients strongly preferred plain-language narratives and simple visuals over dense charts. One participant suggested presenting outputs in narrative form:

> “Make it a narrative form like ‘This person will have a 25% survival rate after three years…’ so that… you’re not just looking at numbers….”

Another participant requested stepwise, page-by-page inputs (“like TurboTax”), with explanations embedded at the point of entry.

Patients also asked that acronyms be spelled out to minimize reliance on a glossary and emphasized the importance of using U.S. customary units:

> “I don’t know how many centimeters I am or how many kilograms I am. Can you make it American standard measurements?”

Providers generally found the interface straightforward and recommended emphasizing allograft survival, incorporating patient-friendly visuals, and ensuring accessibility through web-based and mobile-friendly formats. Some providers suggested that, for select patients, framing outcomes as the probability of remaining on dialysis could reduce anxiety compared with displays focused on “risk of death.”

Coordinators described the format as easy to follow and valued having concrete outputs to support conversations with patients.

#### 3) Functionality: Supportive of clinical judgment; educational value; constraints at the time of offer (phone-based communication and confidentiality)

The app was viewed as a form of decision support that augments, rather than replaces, clinical judgment. Providers emphasized its role as an adjunct to clinical judgment:

> “It’s not going to change your clinical judgment, but it’s going to guide you towards making a better decision,” and “…another tool in that toolbox to provide care to the patient.”

Coordinators anticipated that data-driven output could reassure both providers and patients and support acceptance of some higher-risk offers that might otherwise be bypassed. One coordinator noted that the app “…may actually push providers to accept kidney offers even more…,” particularly when “providers are looking for that extra confirmation.”

Several patients and the patient advocate described the app not only as a decision aid during offer discussions, but also as a self-teaching tool that allowed them to revisit and contextualize transplant risks and benefits outside brief clinical encounters:

> “…with your little app… I understand these numbers… you literally have 10 minutes with your doctor… this actually gives you time to digest everything….”

Providers noted that real-time kidney-offer decisions occur under substantial time pressure and typically occur by phone, which may limit patient access to the app at the point of decision, particularly for those living far from the transplant center.

Donor confidentiality further constrains what patients can see during offer discussions. Detailed donor information cannot be disclosed; patients may be informed if a donor is DCD or high-risk/HCV, but not age, sex, or medical history, limiting patients’ ability to replicate provider inputs in real time. Participants suggested that donor characteristics not available to patients could be represented using ranges or hypothetical values to help contextualize predicted outcomes.

#### 4) Anticipated barriers and facilitators

Anticipated barriers to using the app included inherent clinical uncertainties, digital access limitations, and health literacy challenges. Coordinators emphasized that some clinical events (e.g., primary graft nonfunction) are inherently unpredictable and cannot be fully captured by any predictive model:

> “There’s always going to be those outside factors that you can’t anticipate… nothing is perfect.”

Providers additionally highlighted technology-related barriers and conceptual complexity for some patients, such as difficulties understanding cPRA.

Facilitators included perceived ease of use, willingness to recommend the tool to others, and interest in incorporating it into clinical practice. Providers anticipated greater confidence when considering higher-risk donors, with one noting they might be “…more inclined to take a donor that I probably wouldn’t have using just KDPI.” Providers also described the app’s potential to strengthen patient communication, particularly regarding HCV-positive donors in the direct-acting antiviral era.

In summary, stakeholders emphasized clear risk communication, intuitive stepwise inputs, and a supportive rather than determinative role for decision support within time-sensitive transplant workflows.

## DISCUSSION

Although decision-support tools for DDK transplantation have been described, substantial gaps remain in understanding how transplant stakeholders interpret individualized risk information and how such tools can be effectively integrated into clinical practice.^11,12^ Several themes emerged that have implications for the design and use of kidney-offer decision-support tools. Stakeholders viewed individualized outcome projections as potentially valuable, particularly for higher-risk offers. The themes that emerged highlighted the importance of clinically relevant content, accessible communication, workflow compatibility, and thoughtful implementation.

### Content scope and clinical relevance

Contemporary evidence indicates that transplantation of HCV RNA–positive DDKs into HCV-negative recipients, when paired with timely direct-acting antiviral therapy, yields excellent outcomes.^18–20^ These findings reinforce the need for updated, recipient-directed education that frames HCV-positive donors as viable, evidence-based options for transplantation. Clarifying PHS risk criteria similarly aligns with current practice, including universal donor nucleic acid testing and post-transplant monitoring, and may reduce refusals driven by misperceptions.^21,22^ Although stakeholders raised questions regarding COVID-positive donors, evolving evidence and practice patterns may reduce the relevance of this issue in future iterations of the tool. Content should therefore reflect current evidence, support informed decision making, and complement clinical judgment.

### Health literacy, numeracy, and digital access

Limited health literacy and numeracy are common among transplant candidates and recipients and are associated with worse outcomes and adherence challenges.^23^ Rural and underserved populations also experience persistent telehealth and digital-access gaps, such as lower use of video visits and patient portals, underscoring that patient-directed tools cannot assume uniform internet access or digital skills.^24^ Digital-health equity frameworks emphasize designing tools that accommodate differences in health literacy, technology access, and local resources.^25^ The app’s use of plain-language summaries, U.S. customary units, and printable views aligns with these principles; however, sustained attention to usability for patients with limited connectivity or numeracy will be essential for equitable implementation. These considerations may be especially important for transplant programs serving large rural populations. Stakeholders also described the tool as a resource for learning and reflection outside clinical encounters. These findings are consistent with prior patient-centered design work showing that transplant candidates value concise, easy-to-understand educational content and individualized information to better understand transplant options and likely outcomes.^12^

### Workflow realities: Time pressure and confidentiality

Real-time DDK offer decisions occur under compressed timelines, often by phone, while cold ischemia time continues to accrue, creating pressure for timely acceptance decisions and contributing to the high-stakes nature of offer evaluation.^26^ Strict limits on what donor information can be disclosed to a candidate because of confidentiality requirements often further complicate shared decision making: clinicians may understand the donor–recipient match in more detail, while patients receive only a subset of information.^27^ In this setting, the app may serve as an interpretive aid that translates complex donor and recipient information into understandable, individualized outcome estimates without revealing protected donor details. Reviewing numeric or narrative outputs with patients, even when underlying variables cannot be shared, may support informed decision making and clinician–patient discussions. Consistent with prior work emphasizing integration into existing treatment pathways, stakeholders in our study highlighted workflow compatibility as a prerequisite for adoption, particularly given the time-sensitive nature of DDK offer decisions and the limited donor information that can be shared with patients.^11^

### Implications for design and implementation

Recent appraisal of decision-support tools for DDK transplantation identified shortcomings in usability, actionability, and communication efficacy among existing resources.^11^ Consistent with these observations, participants in our study emphasized that individualized risk prediction alone may be insufficient for successful adoption. Stakeholders viewed the app as a decision-support tool that should augment, rather than replace, clinical judgment and shared decision making.

Several priorities for future refinement emerged. Output displays should prioritize accessible narratives, absolute risks presented as percentages, and optional icon arrays or simple bar charts for patients who benefit from visual aids.^28,29^ Contextual assumptions, such as cold ischemia time ranges and the use of the last versus average donor creatinine, should be transparent and adjustable to allow clinicians to efficiently evaluate plausible offer scenarios. Future versions of the app should reflect current clinical evidence and the realities of transplant practice.

### Limitations

This study reflects perspectives from stakeholders at a single transplant center and was intended to inform refinement of an early prototype. Participants responded to a demonstration of the app rather than using it during actual kidney-offer decisions; therefore, their feedback reflects anticipated use rather than experiences under the time pressure and uncertainty of real-world offer evaluation. Providers were interviewed individually, whereas transplant candidates and coordinators participated in focus groups, which may have influenced the type and depth of feedback obtained. Although reflexive memoing and peer debriefing were used throughout the analysis, involvement of members of the research team in app development may have influenced interpretation of the findings. Because sessions were conducted via Zoom and in English, perspectives of individuals with limited internet access or different language preferences may be underrepresented. Finally, because this study was conducted at a single center, findings may not fully reflect the workflows, patient populations, or practice patterns of other transplant programs. Future work should evaluate decision-support tools during simulated or live kidney-offer encounters across diverse settings.

## Conclusion

Stakeholders viewed the Kidney Risk Calculator app as a promising decision-support tool for kidney-offer discussions. Participants valued individualized donor-recipient risk estimates but emphasized the importance of clear communication, patient education, and compatibility with existing transplant workflows. The app was viewed as a decision aid that should support, rather than replace, clinical judgment and shared decision making.

Future refinements should incorporate updated prediction models, user-friendly displays, and approaches that address differences in internet access, numeracy, and language. Prospective evaluation during simulated or live kidney-offer encounters is needed to assess effects on patient comprehension, clinical communication, acceptance of higher-risk donor kidneys, and organ utilization.

Overall, stakeholders prioritized clear communication and workflow integration alongside individualized risk prediction for kidney-offer decision support.

## Data Availability

All data produced in the present study are available upon reasonable request to the authors

## Data availability statement

De-identified study data may be shared upon reasonable request and execution of a data use agreement. The current version of the prototype web-based Kidney Risk Calculator app is available at https://kcalculator.shinyapps.io/kidneycalc/.

## Acknowledgments

A related abstract based on this work has been accepted for poster presentation at ASN Kidney Week 2026 (Abstract No. 4553088).

## Authors’ contributions

Kelly Chong, PhD (Conceptualization; Funding acquisition; Methodology; Project administration; Supervision; Writing original draft; Review & editing)

Igor Litvinovich, MS (Software; Visualization; Review & editing)

Christos Argyropoulos, MD, PhD (Conceptualization; Supervision; Review & editing)

Rachel Taylor, MFA (Data curation; Formal analysis; Review & editing)

Yiliang Zhu, PhD (Conceptualization; Methodology; Supervision; Review & editing)

## REFERENCES

1. Wolfe RA, Ashby VB, Milford EL, et al. Comparison of mortality in all patients on dialysis, patients on dialysis awaiting transplantation, and recipients of a first cadaveric transplant. N Engl J Med. 1999;341(23):1725–1730. doi:10.1056/NEJM199912023412303

2. Tonelli M, Wiebe N, Knoll G, et al. Systematic Review: Kidney Transplantation Compared With Dialysis in Clinically Relevant Outcomes. Am J Transplant. 2011;11(10):2093–2109. doi:10.1111/j.1600-6143.2011.03686.x

3. Laupacis A, Keown P, Pus N, et al. A study of the quality of life and cost-utility of renal transplantation. Kidney Int. 1996;50(1):235–242. doi:10.1038/ki.1996.307

4. Bae S, Massie AB, Thomas AG, et al. Who can tolerate a marginal kidney? Predicting survival after deceased donor kidney transplant by donor-recipient combination. Am J Transplant Off J Am Soc Transplant Am Soc Transpl Surg. 2019;19(2):425–433. doi:10.1111/ajt.14978

5. Heilman RL, Mathur A, Smith ML, Kaplan B, Reddy KS. Increasing the Use of Kidneys From Unconventional and High-Risk Deceased Donors. Am J Transplant. 2016;16(11):3086–3092. doi:10.1111/ajt.13867

6. Stewart DE, Garcia VC, Rosendale JD, Klassen DK, Carrico BJ. Diagnosing the Decades-Long Rise in the Deceased Donor Kidney Discard Rate in the United States. Transplantation. 2017;101(3):575–587. doi:10.1097/TP.0000000000001539

7. Lentine KL, Smith JM, Lyden GR, et al. OPTN/SRTR 2024 Annual Data Report: Kidney. Am J Transplant. 2026;26(8):S22–S145. doi:10.1016/j.ajt.2026.05.719

8. Rao P, Schaubel D, Guidinger M, et al. A Comprehensive Risk Quantification Score for Deceased Donor Kidneys: The Kidney Donor Risk Index. Transplantation. 2009;88:231–236. doi:10.1097/TP.0b013e3181ac620b

9. Litvinovich I, Ng Y, Chong K, Argyropoulos C, Zhu Y. Predicting Individualized Outcomes for Deceased Kidney Donor Waitlisted Candidates and Recipients. Published online October 2023. doi:10.1101/2023.10.02.23296462

10. Litvinovich I, Chong K, Argyropoulos C, Zhu Y. Prototype web-based Kidney Risk Calculator App. Accessed May 4, 2026. https://kcalculator.shinyapps.io/kidneycalc/

11. White SL, Cutting RB, McLean M, Muscat DM, Patel P, Webster AC. Decision Support Tools in Deceased Donor Kidney Transplantation: An Environmental Scan and Appraisal of Online Resources. Transplantation. 2025;109(12):e682–e690. doi:10.1097/TP.0000000000005473

12. Chu S, Hart A, Bruin M. A patient-centered approach in designing a kidney transplant decision aid. In: 2023. doi:10.54941/ahfe1004234

13. Volk RJ, Llewellyn-Thomas H, Stacey D, Elwyn G. Ten years of the International Patient Decision Aid Standards Collaboration: evolution of the core dimensions for assessing the quality of patient decision aids. BMC Med Inform Decis Mak. 2013;13(S2):S1, 1472-6947-13-S2-S1. doi:10.1186/1472-6947-13-S2-S1

14. Braun V, Clarke V. One size fits all? What counts as quality practice in (reflexive) thematic analysis? Qual Res Psychol. 2021;18(3):328–352. doi:10.1080/14780887.2020.1769238

15. Braun V, Clarke V. Reflecting on Reflexive Thematic Analysis. Qualitative Research in Sport, Exercise and Health. 2019;11(4):589–597.

16. Braun V, Clarke V. Thematic Analysis: A Practical Guide. London, UK: Sage Publications; 2021.

17. NVivo qualitative data analysis software. QSR International Pty Ltd (2020).

18. Schaubel DE, Tran AH, Abt PL, Potluri VS, Goldberg DS, Reese PP. Five-Year Allograft Survival for Recipients of Kidney Transplants From Hepatitis C Virus Infected vs Uninfected Deceased Donors in the Direct-Acting Antiviral Therapy Era. JAMA. 2022;328(11):1102. doi:10.1001/jama.2022.12868

19. Gordon CE, Adam GP, Jadoul M, Martin P, Balk EM. Kidney Transplantation From Hepatitis C Virus–Infected Donors to Uninfected Recipients: A Systematic Review for the KDIGO 2022 Hepatitis C Clinical Practice Guideline Update. Am J Kidney Dis. 2023;82(4):410–418. doi:10.1053/j.ajkd.2022.12.019

20. Sutcliffe S, Ji M, Chang SH, et al. The association of donor hepatitis C virus infection with 3-year kidney transplant outcomes in the era of direct-acting antiviral medications. Am J Transplant. 2023;23(5):629–635. doi:10.1016/j.ajt.2022.11.005

21. Jones JM, Kracalik I, Levi ME, et al. Assessing Solid Organ Donors and Monitoring Transplant Recipients for Human Immunodeficiency Virus, Hepatitis B Virus, and Hepatitis C Virus Infection — U.S. Public Health Service Guideline, 2020. MMWR Recomm Rep. 2020;69(4):1–16. doi:10.15585/mmwr.rr6904a1

22. Organ Procurement and Transplantation Network. Align OPTN Policy with U.S. Public Health Service Guideline, 2020. Rockville (MD): Health Resources and Services Administration; 2021. Accessed March 31, 2026. https://www.hrsa.gov/optn/professionals/resources/patient-safety/optn-policies-to-align-with-2020-us-public-health-service-guideline

23. Pullen LC. A Path Toward Improving Health Literacy and Transplant Outcomes. Am J Transplant. 2019;19(7):1871–1872. doi:10.1111/ajt.15475

24. Rowe Ferrara M, Intinarelli-Shuler G, Chapman SA. Video and Telephone Telehealth Use and Web-Based Patient Portal Activation Among Rural-Dwelling Patients: Retrospective Medical Record Review and Policy Implications. J Med Internet Res. 2025;27:e67226–e67226. doi:10.2196/67226

25. Hatef E, Hudson Scholle S, Buckley B, Weiner JP, Austin JM. Development of an evidence- and consensus-based Digital Healthcare Equity Framework. JAMIA Open. 2024;7(4):ooae136. doi:10.1093/jamiaopen/ooae136

26. McCulloh I, Stewart D, Kiernan K, et al. An experiment on the impact of predictive analytics on kidney offers acceptance decisions. Am J Transplant. 2023;23(7):957–965. doi:10.1016/j.ajt.2023.03.010

27. Organ Procurement and Transplantation Network (OPTN). Guidance for Donor and Recipient Information Sharing. Accessed July 16, 2026. https://www.hrsa.gov/optn/professionals/resources/guidance/guidance-for-donor-and-recipient-information-sharing

28. Ahmed H, Naik G, Willoughby H, Edwards AGK. Communicating risk. BMJ. 2012;344(jun18 1):e3996–e3996. doi:10.1136/bmj.e3996

29. Centers for Disease Control and Prevention (CDC). Numeracy. Published January 31, 2025. Accessed February 22, 2026. Available from: https://www.cdc.Gov/Health-Literacy/Php/Research-Summaries/Numeracy.html.

