## Supplemental Tables S1 & S2 and Figures S1 & S2 for "Personalized Risk-Prediction Tool for Deceased Donor Kidney Offers: Stakeholder Perspectives from a Qualitative Study"

### Supplementary Tables and Figures

#### Supplementary Table S1. Semi-structured interview guide domains and questions

***Learning what users want to see in the App (content)***
What are some initial impressions about the app?
What do you remember most about the app?
Which risk(s) matter the most to you?
Which benefit(s) matter the most to you?
On a scale from 0–10, how much of the information did you understand?
Why did you choose that number?
How can we improve the content to make it more understandable?
What information is missing?
What information would you have liked to know more about to help you make the decision to accept or not accept the kidney offer?

***Learning how users want information displayed in the App (format, features, functionality)***
Now think about the app's format and features; what are your general impressions of the app's display of information?
What features of the app do you find most valuable? And why?
What features do you find unhelpful? And why?
What would you most like to add to or improve about how information is shown on the app?
If you could add or remove one thing from this app, what would it be? And why?

***Understanding user perception of the App***
How would you describe this app to other people?
How likely are you to recommend this app to a friend or colleague?

***Closing***
What are some anticipated barriers and facilitators to adopt this app?
Is there anything we haven't touched on today that you'd like us to know?

**Supplementary Table S2: Quote-Density Matrix (cleaned)**

| Topic | Coordinators facilitators/ maintain | Coordinators limitations/ consider change | Providers facilitators/ maintain | Providers limitations/ consider change | Patients facilitators/ maintain | Patients limitations/ consider change | Total facilitators/ maintain | Total limitations/ consider change |
|---|---|---|---|---|---|---|---|---|
| Overall impressions | 3 | 0 | 4 | 0 | 0 | 0 | 7 | 0 |
| Technical features | 2 | 0 | 1 | 2 | 0 | 1 | 3 | 3 |
| Clarity & usability | 3 | 0 | 4 | 2 | 0 | 3 | 7 | 5 |
| Filling gaps / new / different | 0 | 0 | 2 | 0 | 0 | 0 | 2 | 0 |
| Provider assurance | 3 | 0 | 2 | 0 | 0 | 0 | 5 | 0 |
| Patient communication & education | 3 | 3 | 3 | 5 | 2 | 5 | 8 | 13 |
| High-risk donors (incl. PHS) | 0 | 2 | 2 | 3 | 0 | 0 | 2 | 5 |
| HIPAA / policy | 0 | 0 | 0 | 2 | 0 | 0 | 0 | 2 |
| Patient output visuals | 0 | 0 | 2 | 4 | 0 | 2 | 2 | 6 |
| Transplant characteristics / format | 0 | 0 | 2 | 1 | 0 | 0 | 2 | 1 |
| Age | 0 | 0 | 2 | 0 | 0 | 0 | 2 | 0 |
| Gender | 0 | 0 | 0 | 1 | 0 | 0 | 0 | 1 |
| Height & weight | 0 | 0 | 1 | 1 | 0 | 0 | 1 | 1 |
| Race / ethnicity | 0 | 0 | 1 | 1 | 0 | 0 | 1 | 1 |
| Autoimmune disease | 0 | 0 | 0 | 1 | 0 | 0 | 0 | 1 |
| COPD | 0 | 0 | 0 | 1 | 0 | 0 | 0 | 1 |
| CAD / vascular / HF / DCD | 0 | 0 | 1 | 3 | 0 | 0 | 1 | 3 |
| Diabetes | 0 | 0 | 2 | 2 | 0 | 0 | 2 | 2 |
| Hypertension | 0 | 0 | 1 | 0 | 0 | 0 | 1 | 0 |
| Acceptance time | 0 | 0 | 0 | 1 | 0 | 0 | 0 | 1 |
| Ischemia time (cold/warm) | 0 | 0 | 1 | 3 | 0 | 0 | 1 | 3 |
| COVID-19 impacts | 0 | 2 | 0 | 1 | 0 | 0 | 0 | 3 |
| Creatinine levels | 0 | 0 | 1 | 2 | 0 | 0 | 1 | 2 |
| CVA | 0 | 0 | 0 | 1 | 0 | 0 | 0 | 1 |
| Dialysis | 0 | 0 | 3 | 4 | 0 | 0 | 3 | 4 |
| Donor downtime | 0 | 0 | 0 | 1 | 0 | 0 | 0 | 1 |
| HCV | 0 | 0 | 0 | 3 | 0 | 2 | 0 | 5 |
| Hospitalizations | 0 | 0 | 0 | 1 | 0 | 0 | 0 | 1 |
| Kidney size | 0 | 0 | 0 | 1 | 0 | 0 | 0 | 1 |
| Kidney survival rate | 0 | 0 | 0 | 0 | 0 | 2 | 0 | 2 |
| Medication adherence / compliance | 0 | 0 | 0 | 2 | 0 | 0 | 0 | 2 |
| Number of kidney offers | 0 | 3 | 0 | 0 | 0 | 0 | 0 | 3 |
| Outside factors (unpredictables) | 0 | 2 | 0 | 0 | 0 | 1 | 0 | 3 |
| PRA / CPRA | 0 | 0 | 0 | 2 | 0 | 2 | 0 | 4 |
| Pre-transplant transfusions | 0 | 0 | 0 | 1 | 0 | 0 | 0 | 1 |
| Smoking / alcohol / substance use | 0 | 0 | 0 | 2 | 0 | 0 | 0 | 2 |
| Urine output | 0 | 0 | 0 | 1 | 0 | 0 | 0 | 1 |

Aggregated counts of coded references by topic, stakeholder group (coordinators, providers, patients), and valence (facilitators/maintain vs. limitations/consider change) were generated using NVivo matrix queries to examine cross-group representation. Counts are descriptive only and were used to support quotation selection and coverage checks; they were not used to weight themes, consistent with reflexive thematic analysis.

**Supplementary Figure S1**. Top 10 Topics by Limitation/Consider Change (Quote-Density Heatmap).

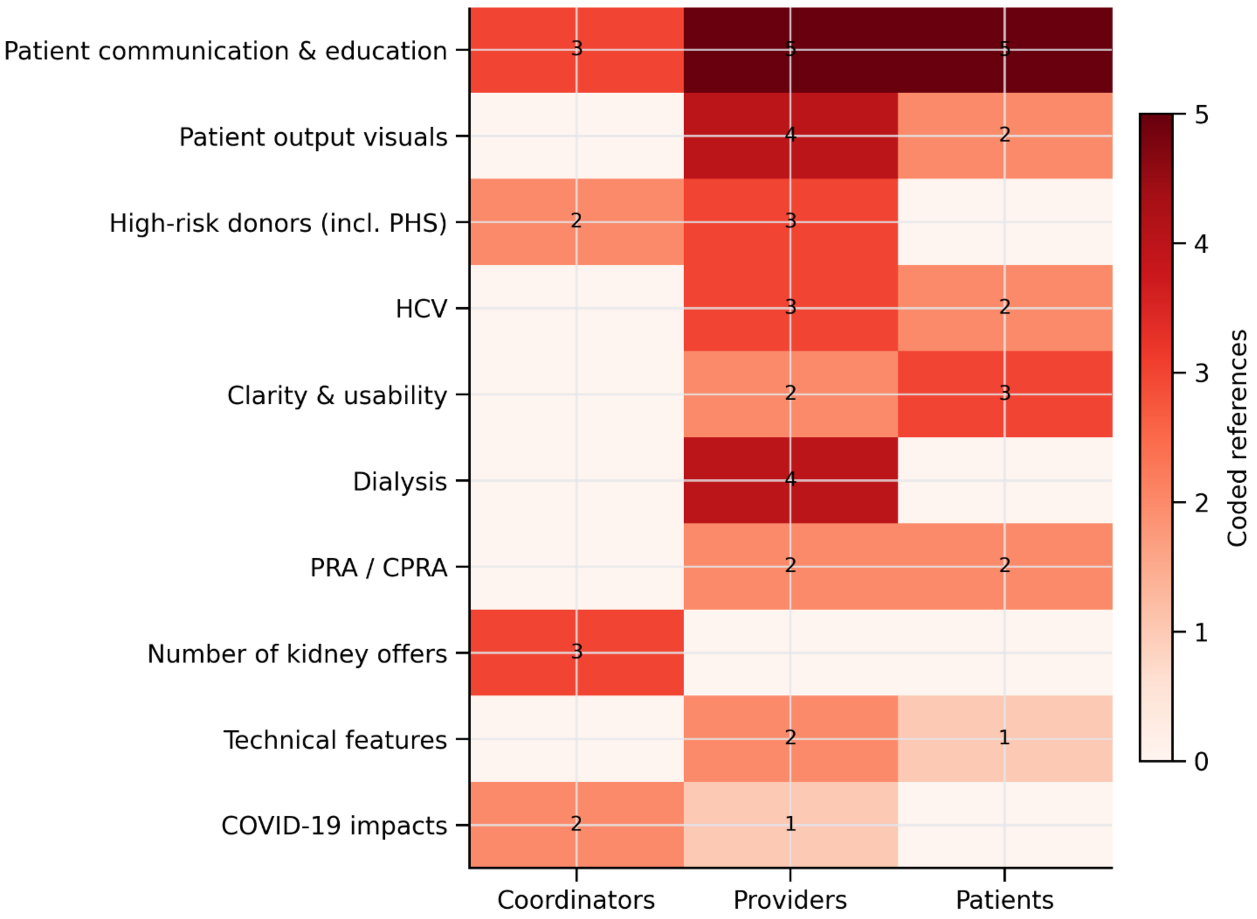


Rows list the top 10 topics by coded-reference density for each valence; columns are coordinators, providers, and patients. Cell labels show coded references per topic-by-group combination; color intensity reflects magnitude.

**Supplementary Figure S2. Top 10 Topics by Facilitators/Maintain (Quote-Density Heatmap).**

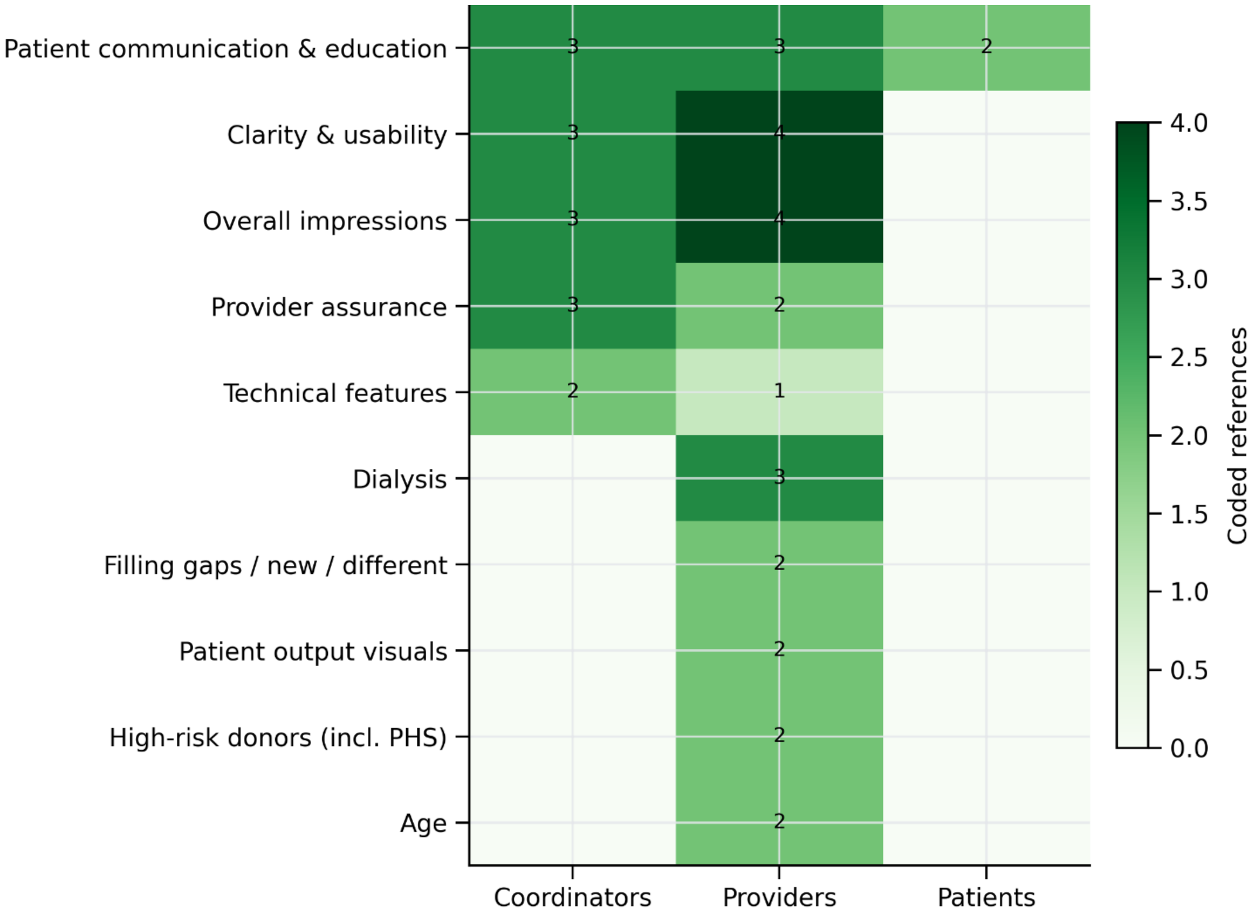


**Disclosure:** Heatmaps were generated from the cleaned quote-density matrix using AI-assisted code generation (Microsoft 365 Copilot with enterprise data protection) to automate plotting. All labels, counts, and figure outputs were reviewed and verified by the authors. The author used the following prompt:

> "Generate two separate heatmap panels from the attached "combined matrix" excel file—one for Limitations / Consider change (in red) and one for Facilitators / Maintain (in green). Show only the top 10 topics for each valence based on total coded references across coordinators, providers, and patients. Plot groups on x-axis and topics (no IDs) on y-axis. Show model output."

**Reproducibility note:** Python code is available from the corresponding author upon reasonable request.
